# High-Resolution Spatial Transcriptomics Reveals Interferon-Associated Immune Niches and Antigen Presentation Programs in Inclusion Body Myositis

**DOI:** 10.64898/2026.08.23.26361142

**Authors:** Sandra de Haan, Caro van Andel, Laura Heezen, Ramon Arens, Hermien E. Kan, Umesh Badrising, Ahmed Mahfouz, Pietro Spitali

## Abstract

Inclusion body myositis (IBM) is a progressive inflammatory myopathy characterized by muscle fiber degeneration, immune infiltration, and protein aggregation. Despite the prominent immune infiltrates that characterizes IBM muscle, the factors driving immune infiltration remain unknown, and the repertoire and spatial organization of infiltrating immune populations remain poorly defined. Here, we used high-resolution spatial transcriptomic profiling to define the cellular and spatial architecture of IBM muscle. Immune profiling revealed a complex inflammatory landscape dominated by interferon-responsive CD8+ T cells and interferon-stimulated antigen-presenting macrophages, which organized into spatially localized immune hubs surrounding myofibers. Myofibers within these immune-rich microenvironments exhibited increased expression of interferon-responsive genes and HLA class I and II antigen presentation machinery components across fiber subtypes. In addition, we identified muscle-intrinsic remodeling and regenerative programs that may precede or contribute to immune recruitment, characterized by focal spatial activation of genes involved in proteostasis, cytoskeletal organization, and myofiber repair. Together, these findings define the spatial immune landscape of IBM muscle and reveal coordinated immune and muscle-intrinsic programs that shape disease pathology.

## Introduction

Inclusion body myositis (IBM) is the most common acquired inflammatory myopathy in older adults and is characterized by progressive muscle weakness, myofiber degeneration, immune infiltration, and accumulation of protein aggregates [1, 2]. Despite its inflammatory features, conventional immunosuppressive therapies provide limited clinical benefit, underscoring the need to understand the mechanisms that sustain immune-mediated muscle injury. Several targeted treatments are currently being studied, including approaches that target KLRG-1-positive, highly differentiated cytotoxic T cells (Ulviprubart), selectively inhibit T-effector cells while preserving regulatory T cells (Sirolimus), or block interferon signaling pathways (Ruxolitinib) [3–5]. Immune profiling of peripheral blood and immunological studies of muscle have not resulted in a unifying pathogenic concept explaining how inflammation is initiated and sustained in IBM. Defining immune–myofiber interactions is therefore essential for understanding the mechanisms initiating and sustaining IBM pathology, for guiding the development of targeted therapies, and for providing insight into why current therapeutic approaches have limited success.

Recent advances in high-resolution spatial transcriptomic technologies enable characterization of complex tissue microenvironments while preserving cellular spatial relationships. These approaches provide an opportunity to resolve the composition and molecular states of immune infiltrates in IBM muscle and identify the transcriptional programs of myofibers associated with immune activity.

In this study, we established a high-resolution spatial map of the IBM muscle microenvironment to define the cellular organization underlying disease pathology. We identified spatially organized immune hubs composed of interferon-responsive CD8+ T cells and macrophages with an increased capacity for antigen presentation that co-localize around specific myofibers. These immune-associated myofibers exhibited activation of interferon signaling and antigen presentation programs across fiber subtypes. We additionally identified muscle-intrinsic remodeling and regenerative programs that may precede immune infiltration, characterized by genes involved in proteostasis and cytoskeletal remodeling with distinct focal spatial expression patterns. Together, these findings reveal a spatially organized immune–myofiber axis in IBM and thereby provide insight into the molecular features of muscle fibers associated with persistent inflammatory activation.

## Results

### Experimental set-up and data characterization

To define the cellular and spatial architecture of inclusion body myositis (IBM) muscle, we performed Xenium spatial transcriptomics on three IBM patient biopsies and three healthy control (HC) biopsies. Serial sections from each biopsy were analyzed using two complementary gene panels: (1) the pre-configured 5,000 Gene Human Pan-Tissue & Pathway Panel (hereafter 5K panel), designed to capture broad cellular states through genes encoding receptors, ligands, and signaling molecules; and (2) a custom 480-gene panel (hereafter 480 panel), enriched for muscle-specific genes, inflammatory pathways, antigen presentation markers, and neuromuscular disease-associated genes to enable detailed muscle fiber subtype annotation. In addition to their complementary gene content, both panels contained overlapping canonical marker genes, enabling robust annotation of the major cell types in each dataset (**Supplement Table 1**).

Cell segmentation was performed independently for muscle fibers and mononucleated cells. Across all samples, this resulted in the identification of 10,812 and 11,014 segmented muscle fibers in the 5K and 480 panels, respectively, alongside 151,485 and 144,355 mononucleated cells. Leveraging the serial-section experimental design, segmented muscle fibers were matched across panels, enabling integration of gene expression data from the 5K and 480 panels, resulting in 8,998 matched fibers (**Fig. 1A–B**). Clustering of all segmented cells enabled the assignment of broad cell identities across the muscle tissue. Based on canonical marker genes, we identified all major cellular compartments, including myofibers/myonuclei (*MYH1^+^*/ *MYH7^+^*), satellite cells (*PAX7^+^*), immune cells (*CD8A^+^*/ *CD4^+^*/ *CD79A^+^*), fibro-adipogenic and collagen-producing cells (*COL12A1^+^*), adipocytes (*LPL^+^*), and vascular cells including endothelial cells (*VWF^+^*) and pericytes (*RGS5^+^*) (**Fig. 1C**). Comparison of cell type composition between IBM and HC biopsies revealed a marked expansion of immune cell populations in IBM, consistent with the characteristic inflammatory infiltrates of the disease (**Fig. 1D**). To visualize the spatial distribution of the identified cell types, annotated cell identities were mapped back to their original tissue coordinates (**Fig. 1E**).

**Fig 1:**
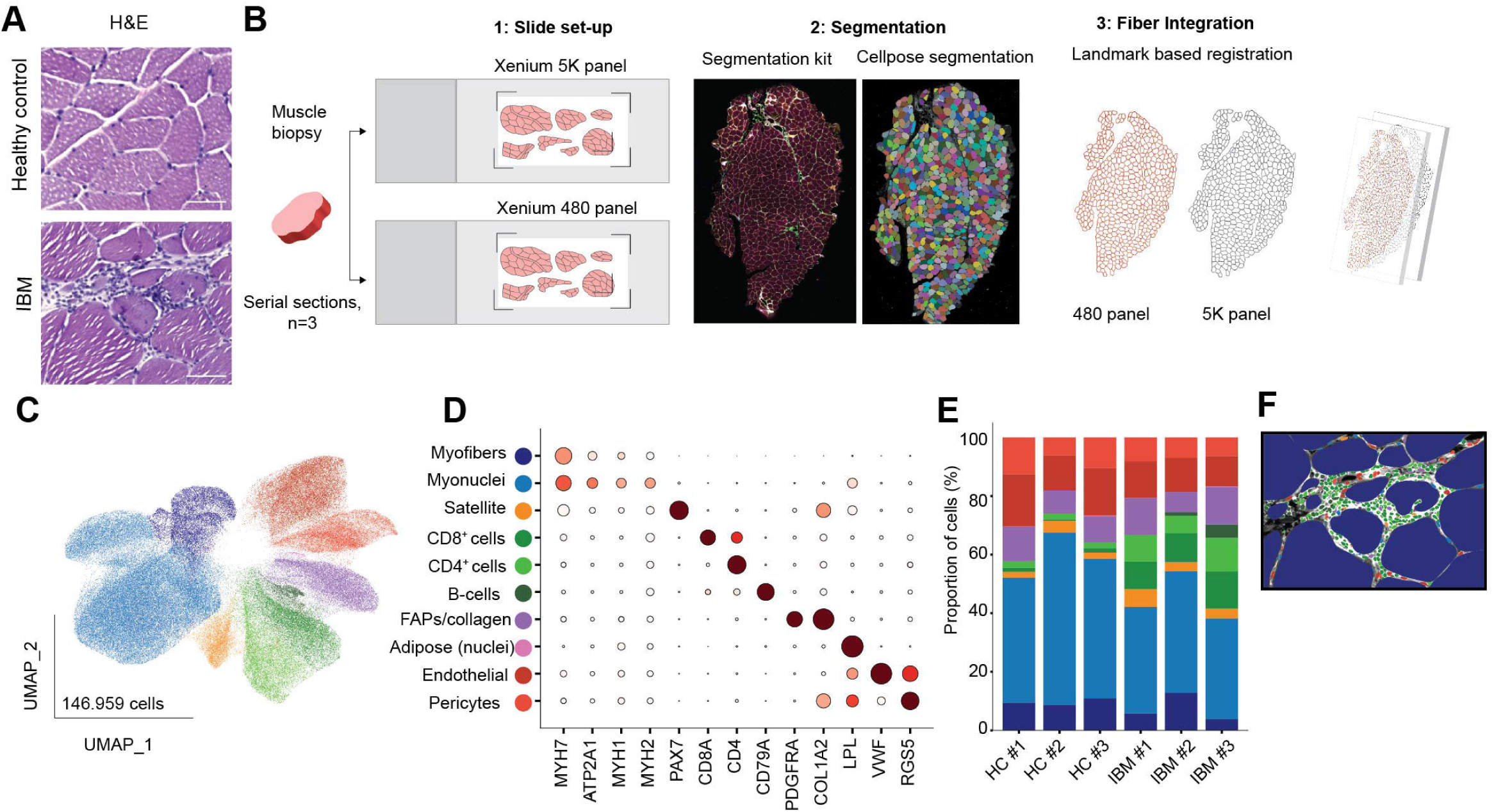
Overview of experimental set-up and IBM histology. **A)** H&E image of healthy control (top), and IBM (bottom) muscle biopsies. **B)** Sketch showing experimental design, slides set-up and analysis pipeline. **C)** UMAP visualization of all segmented object in 480 panel dataset. **D)** Dotplot showing markers genes of identified cell types/clusters. **E)** Compositional analysis of spatial transcriptomics data of HC and IBM biopsies, per sample showing an increase in immune cell populations. **f)** Representative image of segmented objects color coded for cell types identified of IBM biopsy example as shown in A. FAPs, fibroadipogenic progenitors; HC, healthy control; IBM, inclusion body myositis.

### Selective type 2 fiber loss and emergence of regenerative and damaged myofibers in IBM

For in-depth characterization of the myogenic compartment myofibers were analyzed using the integrated dataset. Subclustering resolved distinct myofiber populations, including canonical type 1 (*MYH7+/MYH7B+*), type 2a (*ATP2A1+/MYH1+*), and type 2a (*ATP2A1+/MYH2+*) myofibers, which were present in both HC and IBM muscle (**Fig. 2A-B**). In contrast, IBM muscle contained an early regenerative population, characterized by the expression of *MYH3*, and a late regenerative population characterized by the expression of *MYH3* and *MYH8*. Additionally, a fiber population characterized by the reduced expression of canonical myofiber subtype markers (*MYH7/MYH1/MYH2*), and the expression of *SAA1-4*, was classified as damaged. While healthy muscle displayed an approximately balanced distribution of type 1 and type 2 fibers, IBM biopsies showed a marked predominance of type 1 fibers, reflecting a preferential depletion of type 2 fiber populations, accompanied by the emerge of regenerative and damaged myofiber populations (**Fig. 2C**).

**Fig. 2:**
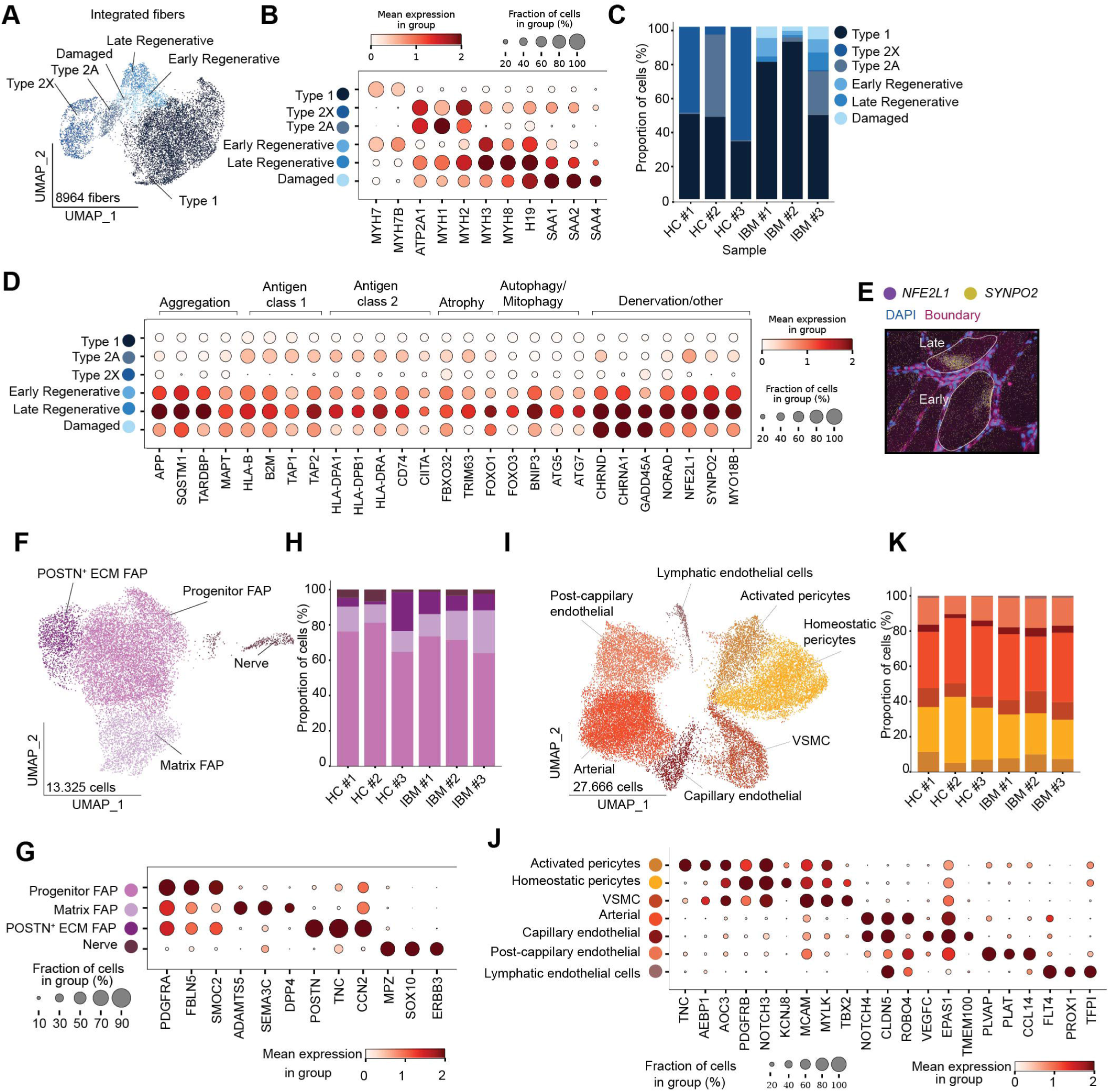
Subtype analysis of myocyte compartment, FAPs and vascular cells. **A)** UMAP visualization of myofiber populations. **B)** Dotplot showing markers genes for the different myofiber populations of A**. C)** Compositional analysis of myofibers showing selective loss of type 2 fibers in IBM, and the emerge of early and late regenerating and damaged myofiber subtypes. **D)** Dotplot showing markers genes of disease related transcripts. **E)** Spatial visualization of focal expression of *NFE2L1* and *SYNPO2* in early and late regenerative fibers. **F)** UMAP visualization of sub clustering of FAPs. **G**) Dotplot showing markers genes for identified FAP clusters. **H**) Compositional analysis of FAPs, showing no major changes in cell type composition. **I)** UMAP visualization of sub clustering of vascular cells. **J)** Dotplot showing markers genes for identified vascular cell types. **K)** Compositional analysis of vascular cells, showing different population of vascular cells. FAPs, fibroadipogenic progenitors; HC, healthy control; IBM, inclusion body myositis, VSMC; vascular smooth muscle cells; ECM, extracellular matrix.

To further characterize the regenerating and damaged populations, expression of previously identified disease-related transcripts was assessed. Expression of transcripts of known protein aggregates described in IBM, including amyloid beta precursor protein (*APP*), sequestosome 1 (*SQSTM1, p62*), TAR DNA-binding protein 43 (*TARDBP, TDP-43*), and microtubule-associated protein tau (*MAPT, tau*), as well as MHC-class II (*HLA-DPA1, HLA-DPB1, HLA-DRA, CD74, CIITA*), were predominantly expressed in late regenerative fibers, and to a lower extend in early regenerating fibers and damaged fibers. Fibers assigned as early or late regenerative fibers were additionally enriched for atrophy and autophagy related markers (*TRIM63/BNIP3*). Recent studies have identified *GADD45A* as a marker of damaged myofibers [6]. In our dataset, *GADD45A* expression was detected in both damaged and late regenerative fiber populations, and to a lower extend early regenerating fibers, suggesting activation of shared cellular programs in these myofiber populations. These populations also showed increased expression of the stress-associated long non-coding RNA N*ORAD*, which has been implicated in functional denervation responses. Consistent with a denervation-associated state, regenerative and damaged myofibers exhibited enrichment of neuromuscular junction-associated genes, including *CHRND* and *CHRNA1* (**Fig2. D**). Differential expression analysis identified genes associated with early and late regenerative fiber states (**Supplementary Table 2**). Subsequent visual inspection of spatial expression patterns revealed a subset of markers, including *NFE2L1* (Nuclear Factor, Erythroid 2 Like 1), *SYNPO2* (Synaptopodin 2), and *MYO18B* (Myosin 18B), that showed focal enrichment within discrete regions of regenerative fibers (**Fig. 2E**). Functionally, these genes are associated with proteasome activity, protein quality control, and cytoskeletal remodeling, and their enrichment may reflect activation of cellular programs involved in regenerative myofiber states.

Of note, a subset of type 2a and type 1 fibers also displayed focal enrichment of *NFE2L1* and *SYNPO2*, but lacked *MYH3* and *MYH8* expression, suggesting these fibers may represent an early remodeling/regenerative state preceding detectable activation of the regenerative program (data not shown).

### Preserved fibro-adipogenic progenitor and vascular cellular composition in IBM muscle

Given the involvement of multiple cell types in IBM tissue remodeling, we sought to determine the cellular composition and transcriptional states of non-myofiber compartments within IBM muscle. Subclustering of the fibro-adipogenic progenitor (FAP)/extracellular matrix (ECM)-associated compartment identified four major populations: progenitor FAPs, ECM-producing FAPs, matrix-remodeling FAPs, and a nerve-associated Schwann cell population (**Fig2.E-F**). The relative abundance of these populations was comparable between HC and IBM samples, suggesting that major alterations in FAP composition were not a dominant feature of IBM muscle at the transcriptional population level (**Fig.2 G**).

Similarly, subclustering of the vascular compartment identified distinct vascular populations, including activated and homeostatic pericytes, vascular smooth muscle cells (VSMC), arterial endothelial cells, capillary endothelial cells, post-capillary endothelial cells, and lymphatic endothelial cells (**Fig 2.H-I**). These vascular subpopulations were detected in both HC and IBM samples with broadly similar relative proportions, indicating preservation of overall vascular cellular composition despite the extensive myofiber pathology observed in IBM (**Fig 2.J**).

### IBM exhibits a diverse, IFN-associated immune microenvironment

To date, the immune repertoire in IBM has not been extensively characterized. We therefore performed immune-cell subclustering using the 5K panel, which was selected for its broader transcriptomic coverage and signaling molecules relevant to immune-cell identity and activation state. This enabled the identification of various T-cell populations, including CD4+ T-cells (*CD4^+^/CD3E^+^*), regulatory T-cells (*CD4^+^/CD3E^+^/FOX3P^+^*), cytotoxic CD8+ T-cells (*CD3E^+^/ CD8A^+^/ GZMH^+^*), IFN-stimulated cytotoxic T-cells (*CD3E^+^/ CD8A^+^/ GZMH^+^/ STAT1^+^/ GBP5^+^/ CXCL9^+^),* proliferative CD8+ T-cells (*CD3E^+^/ CD8A^+^/ MKI67^+^*), Terminal effector T-cells (*CD3E^+^/CD8A^+^/ GZMB^+^/ PRF1^+^*), as well as B-cells (*CD79A^+^*), and plasma cells (*XBP1^+^/ CD79A^+^*) (**Fig 3A-B)**. In line with a recent study reporting plasma cell transcript localization patterns [7], spatial visualization of plasma cells revealed that plasma-cell-associated immunoglobulin transcripts, including *IGHG1* and *IGHG4*, was found outside of the plasma cell nuclear boundaries and displayed a circular distribution surrounding plasma-cell nuclei, consistent with the high abundance of immunoglobulin transcripts characteristic of antibody-producing plasma cells (**Supplement Fig. 1**).

**Fig. 3:**
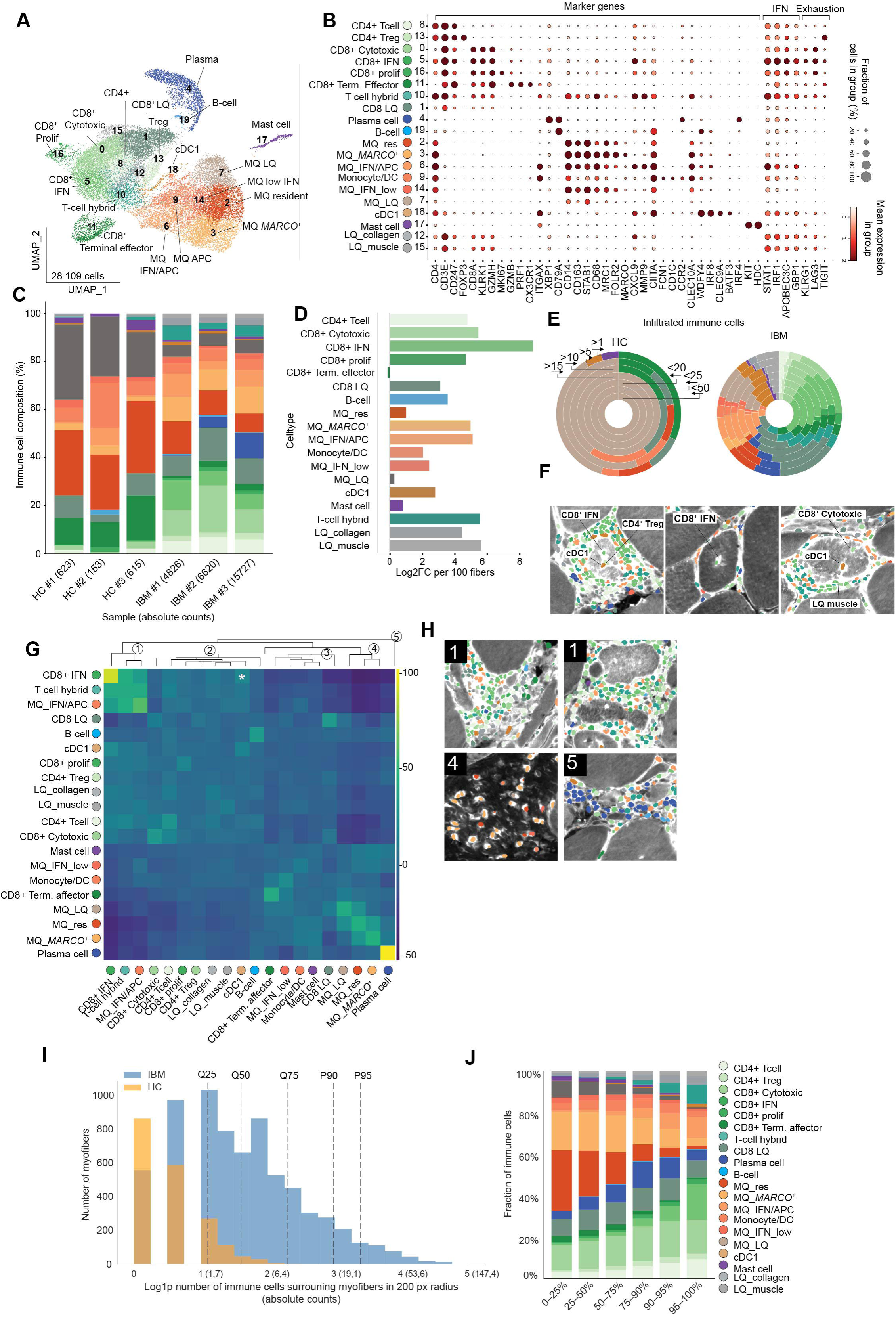
Complex immune repertoire with colocalization of IFN stimulated CD8+ T-cell and antigen-presenting macrophages at advance stages of disease. **A)** UMAP visualization immune cells of the 5000 panel. **B)** Dotplot showing markers genes for celltypes identified in A. **C)** Compositional analysis of immune cells per sample, showing an increase in IFN+ responsive CD8+ T-cells and antigen presenting macrophages. **D)** Horizontal bar plot showing log2 fold changes in immune cell abundance normalized per 100 myofibers for IBM compared to HC for different immune subtypes. **E)** Donut plot showing the composition of immune cells at increasing inward pixel distances from the myofibers membrane. **F)** Representative spatial maps showing annotated immune cell populations of infiltrated immune cells. **G)** Neighborhood enrichment heatmap with hierarchical clustering, scale bar showing neighborhood enrichment score z-scores. **H**) Representative spatial maps of branches identified in G. **I)** Histogram of log-normalized total immune cells surrounding myofibers in a 200 pixel radius. **J)** Stacked bar plot of immune cell composition across immune load quartiles/percentiles of immune positive cells. LQ, low quality; MQ, macrophage; IFN, interferon; APC, antigen-presenting; DC, dendritic cell; cDC, conventional dendritic cell; Q, quantile, P, percentile; px, pixel.

The IFN-responsive cytotoxic CD8+ T-cell population displayed expression of pan-interferon markers (*STAT1, IRF1*), together with type I interferon-associated response genes including *APOBEC3C* and type II interferon-associated genes such as *GBP1*. In addition, the inhibitory receptors *LAG3* and *TIGIT*, which have been associated with chronic T-cell activation and exhaustion [8] were predominantly expressed by CD8+ cytotoxic and IFN-stimulated CD8+ T-cell populations (**Fig.3B**).

*KLRG-1*, a cell-surface receptor associated with highly differentiated cytotoxic T cells and a potential therapeutic target for depletion of this effector population [9], was predominantly expressed by T-cell populations. The highest proportion of *KLRG-1*-positive cells was observed among CD8+ IFN-stimulated T cells (38.98%) and CD8+ cytotoxic T cells (34.54%), whereas the remaining T-cell populations showed *KLRG-1* positivity in fewer than 20% of cells (**Fig. 3B**, **Supplement Fig. 2**).

Different myeloid populations included resident macrophages (*CD14^+^/CD163^+^/CD68^+^/MRC1^+^/FOLR2^+^*), MARCO^+^ macrophages (*CD14^+^/CD163^+^/CD68^+^/MARCO^+^*), IFN-stimulated macrophages (*CD14^+^/CD68^+^/STAT1^+^/CXCL9^+^/MMP9^+^*), a monocyte/DC interface compartment (*CD14^+^/CCR2^+^/CLEC10A^+^*), a moderately IFN-stimulated macrophage population (*CD68^+^/MRC1^+^/STAT1^+low^*), dendritic cells type 1 (*CLEC9A^+^/WDFY4^+^/IRF8^+^*) and mast cells (*KIT^+^/HDC^+^*). The IFN-stimulated, antigen-machinery enriched macrophage population was characterized by expression of *STAT1* and *CXCL9*, indicative of a type II interferon activation. Additionally, a population with concurrent T-cell and macrophage marker expression was observed in immune-dense regions. Given the close proximity of lymphoid and myeloid cells, these mixed profiles may reflect segmentation overlap, transcript mixing, or phagocytic uptake of cellular material. This population was therefore classified as a ‘T-cell hybrid’ population (**Fig 3A-B, Supplement Fig.3, Supplement Table 3**).

In HC biopsies, the immune repertoire mainly consists of resident macrophages, whereas IBM biopsies presented a more diverse immune repertoire characterized predominantly by expansion of various T-cell populations. In contrast to IBM, HC samples lacked regulatory T-cells and plasma cells (**Fig. 3C**). Next, we normalized immune cell abundance to the number of myofibers per biopsy and observed an increase in immune cell density from 100,6 ± 68.7 (mean ± SD) immune cells per 100 myofibers in HC to 400.6 ± 308.4 immune cells per 100 myofibers in IBM (**Supplement Fig. 4**). This expansion was primarily driven by IFN-responsive CD8 T cells (log2 fold change = 8.2), followed by increases in multiple other T-cell subsets, MARCO+ macrophages, and IFN-stimulated antigen-presenting macrophages (**Fig. 3D**). Despite the overall expansion of the T-cell compartment, CD8+ Terminal effector T-cells exhibited a negative log2 fold change in IBM relative to HC, indicating a relative reduction of this terminally differentiated cytotoxic T-cell population per myofiber, suggesting that chronic inflammatory signaling in IBM may be associated with altered or impaired progression toward a terminal effector cytotoxic state.

As immune involvement in IBM extends beyond endomysial immune cell accumulation surrounding fibers, and is additionally characterized by immune cells invading non-necrotic myofibers, we leveraged the spatial resolution of our dataset to identify and characterize immune nuclei detected within myofiber segmentation masks. We observed that infiltrating immune cells comprised multiple immune cell populations, including cytotoxic T cells, CD8⁺ IFN-responsive T-cells, cDC1 cells, CD4⁺ Treg cells, and macrophages. As different immune populations may have distinct roles in IBM pathology, we assessed whether immune subsets exhibited preferential localization near the myofiber membrane or were detected at greater inward distances by assessing the immune composition at various inward buffer distances from the myofiber membrane. The cellular composition of infiltrated immune cells was largely maintained across increasing inward distances from the myofiber membrane, although macrophages were more frequently detected close to the membrane, whereas T-cell and cDC1 populations remained represented at greater inward distances, centrally located within individual myofibers (**Fig. 3E-F**).

### Increasing immune load reflects a transition toward IFN-associated inflammatory hubs in IBM

As immune cells in IBM muscle preferentially accumulate around specific myofibers, we next asked which immune populations colocalize more or less frequently than expected by chance by performing neighborhood enrichment analysis (**Fig. 3G**). Hierarchical clustering of neighborhood enrichment scores identified five distinct spatial immune neighborhoods within IBM muscle.

The strongest colocalization was observed between IFN-stimulated antigen-presenting macrophages, IFN-responsive CD8 T cells, and the T-cell hybrid cluster, indicating a highly organized IFN-associated immune niche (**Fig. 3G–H**). A second neighborhood comprised the majority of lymphocyte populations, including cytotoxic CD8 T cells, CD4 T cells, regulatory T cells, proliferating CD8 T cells, cDC1 cells, B cells, together with low-quality muscle and collagen-associated immune cells, which showed moderate levels of enrichment. It is interesting to note that cDC1 cells, cells that have previously been proposed as interacting partners of KLRG-1+ CD8 T cells based on their expression of KLRG-1 ligands Cadherin-1 (CDH1) and Cadherin-2 (CDH2) [10], did segregate into different hierarchical branches based on their overall spatial interaction profiles. However, their pairwise enrichment indicates that these populations frequently co-localize within IBM muscle (**Fig. 3G, asterisk**). Supporting this potential interaction, cDC1 cells were the main population expressing *CDH1* and *CDH2* in our dataset (**Supplementary Fig. 5**), suggesting that cDC1 cells may represent an interacting component of the IFN-associated immune microenvironment.

A third neighborhood consisted of terminal effector CD8 T cells, cells of the monocyte/DC interface, moderately IFN-stimulated macrophages, and mast cells. Within this group, terminal effector CD8 T cells showed stronger colocalization with monocytes and macrophages than with other T-cell populations. Notably, because terminal effector CD8 T cells were predominantly detected in healthy control tissue, their association with monocytes and macrophages in IBM biopsies may reflect homeostatic immune surveillance rather than IBM-specific pathology. A fourth neighborhood was formed by *MARCO*+ macrophages, resident macrophages, and low-quality macrophages. Although these populations were distributed throughout the biopsy, their strongest colocalization occurred within the perimysium (**Fig. 3G–H**). Finally, plasma cells were present throughout the biopsy, and showed localized enrichment and preferential colocalization with each other, forming distinct plasma cell neighborhoods (**Fig. 3G-H**).

Next, we sought to study how immune cell composition surrounding myofibers changes with increasing immune cell load within a 200-pixel radius around each myofiber (**Fig. 3I, Supplement Fig. 6**). This measure of local immune-cell abundance per myofiber is hereafter referred to as immune load per myofiber. Myofibers exhibiting the highest immune load were surrounded by dense immune-cell accumulation, referred to as immune hubs of immune cells. At the lowest immune load percentile, surrounding immune hubs where composed predominantly of resident and *MARCO+* macrophages, whereas with increasing immune load, composition changes to IFN-responsive CD8+ T cells as well as IFN stimulated antigen presenting macrophages, and hybrid T-cells **(Fig. 3J**). Together, these results suggest that IBM progression is associated with the emergence of spatially organized IFN-associated immune hubs surrounding myofibers.

Given the prominent expansion and spatial association of IFN-responsive CD8+ T cells within these inflammatory hubs, we next sought to identify cell-surface markers that characterize this population and may, similar to KLRG-1, represent potential targets for selective modulation or depletion. We therefore assessed the percentage of cells expressing candidate genes in the CD8+ IFN-stimulated T-cell population and compared their expression across other T-cell populations and other immune populations. Markers were selected based on enrichment in CD8+ IFN-stimulated T-cells (>30% of cells), while being expressed in no more than 30% of cells in the non-CD8+ T-cell populations, identifying 105 candidate genes (**Supplementary Table 4**). Among these, the IL-21 receptor (*IL21R*), chemokine receptor *CCR5*, SLAM-family lymphocyte surface receptor *LY9* (*CD229*), and Fas ligand (*FASLG*) were identified as candidate cell-surface markers enriched in the CD8+ IFN-responsive T-cell population. Notably, *FASLG* displayed an expression pattern resembling that of *KLRG-1* **(Supplementary Fig. 7**).

### Immune-enriched myofibers exhibit type I- and type II interferon activation and antigen presentation

Having identified IFN-associated immune hubs surrounding myofibers, we next investigated whether specific myofiber populations were preferentially associated with these hubs, as previously quantified within a 200-pixel radius around each myofiber. Significant differences in immune cell load for different myofiber populations were observed (Kruskal–Wallis test, H = 347.56, p = 5.87 × 10⁻□³). Post-hoc Dunn’s testing with Benjamini–Hochberg false discovery rate (FDR) correction revealed significantly increased immune burden surrounding late and early regenerating fibers compared with canonical fiber populations (FDR-adjusted p < 0.001), with late regenerating fibers displaying the highest immune cell load, followed by early regenerating fibers.

To identify transcriptional programs associated with increased immune load, we used linear regression to identify transcripts expressed in myofibers whose expression correlated with increasing immune cell abundance surrounding myofibers (**Fig.4C**). Genes positively associated with immune abundance surrounding myofibers included pan-interferon transcripts including tryptophanyl-tRNA synthetase (*WARS),* Interferon-Induced Protein 35 (*IFI35*), Interferon Regulatory Factor 1 (*IRF1*). Notably, WARS belongs to the aminoacyl-tRNA synthetase (aaRS) family, several members of which have been implicated as autoantigens in other idiopathic inflammatory myopathies, but its biological significance in IBM remains largely unexplored. In addition to pan-IFN markers, markers of both an type I and an type II interferon were associated with an increase immune burden, including *IFIT3*, *APOL1*, *APOL2* (type I), and *GBP1*, *CXCL9* (type II), respectively. In addition to IFN-inducible genes, transcripts associated with antigen presentation, including HLA class II molecules, as well as components of the antigen presentation machinery, including *TAP1* and *TAP2*, were associated with an increase in immune cells surrounding myofibers, indicating activation of the antigen presentation machinery and possible antigen presentation (for full list see **Supplement Table 5**). To account for differences in myofiber subtype composition, the analysis was repeated independently for each myofiber population, revealing a conserved transcriptional response across fiber subtypes, dominated by interferon signaling and antigen presentation pathways (**Fig.4D-E, Supplement Table 6**). Strikingly, zonated expression of pan, type I and type II IFN transcripts was observed at sites of moderate immune infiltration (**Fig.4D-E, left panels)**. Together, these findings indicate that immune-enriched myofibers exhibit a broad interferon-associated transcriptional program involving both type I- and type II interferon-associated signatures, together with enhanced expression of antigen presentation pathways, suggesting an increased capacity for antigen processing and presentation.

**Fig. 4:**
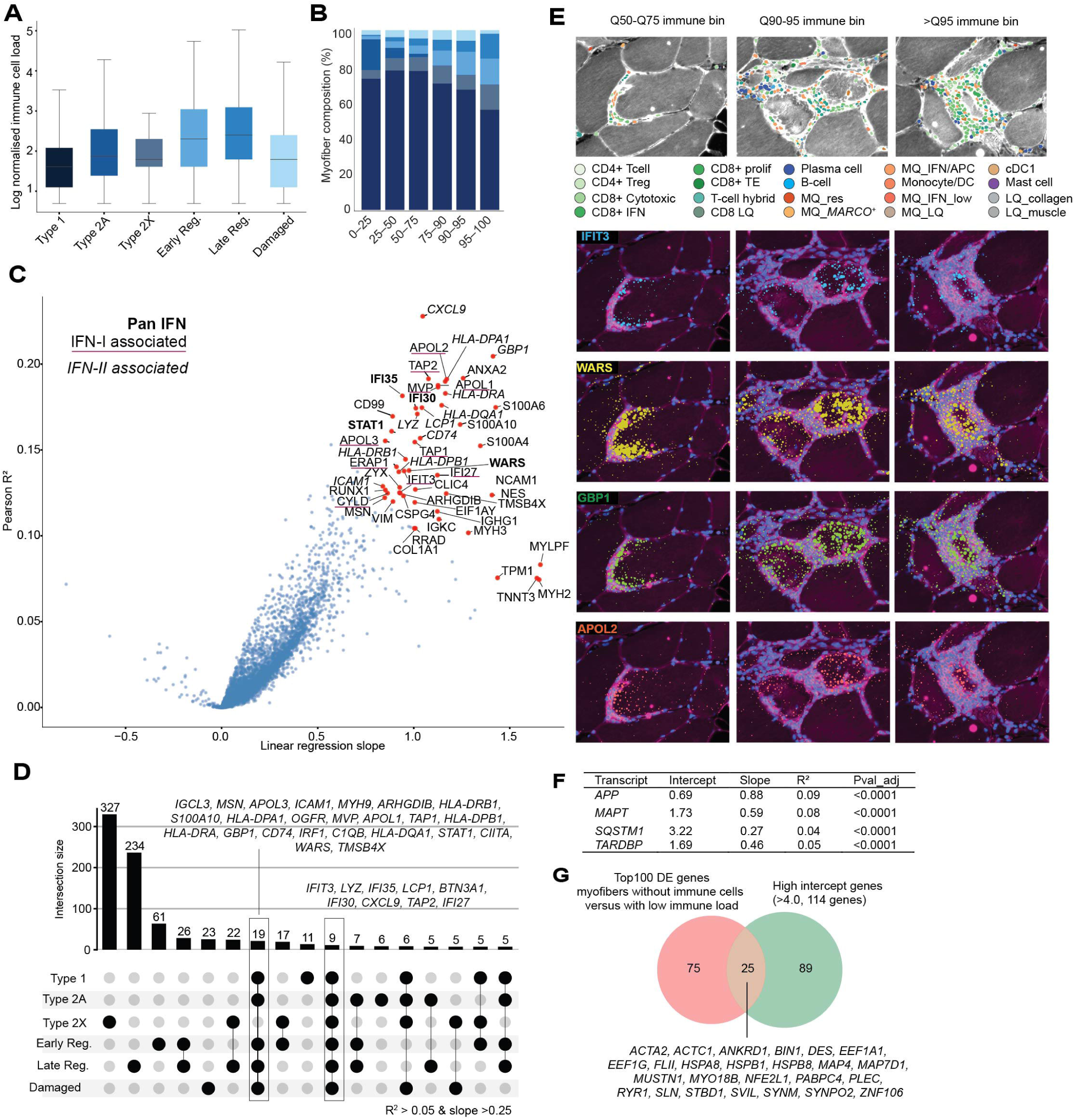
Immune enriched myofibers present enrichment of interferon-responsive and antigen presentation related transcripts. **A)** Boxplot showing log normalized total immune cells across myofiber populations. **B)** Stacked bar plot showing the composition of myofiber populations across increasing immune burden percentiles (0–25, 25-50%, 50–75%, 75–90%, 90–95%, and >95%). **C)** Scatter plot of linear regression results highlighting genes which expression increase with increased immune load surrounding myofibers. The x-axis represents the regression slope, and the y-axis the coefficient (R²), with the top 50 genes highlighted. **D)** UpSet plot showing overlap of transcripts associated with increasing immune load across individual myofiber populations. Bars indicate the number of shared or unique genes, demonstrating a conserved interferon- and antigen presentation-associated transcriptional response across fiber subtypes. **E)** Representative spatial visualization of expression of transcripts within myofibers with increasing local immune burden, showing expression of interferon-responsive transcripts identified in **C. F**) Linear regression results for transcripts of proteins known to aggregate in IBM. **G**) Venn diagram showing the overlap between the top 100 DE genes identified in myofibers with low immune load compared to no immune load and the 114 high intercept genes (>4.0) identified through linear regression analysis. IBM, inclusion body myositis; HC, healthy control; IFN, interferon; APC, antigen-presenting cell; HLA, human leukocyte antigen; MHC, major histocompatibility complex.

Next, we studied how expression of transcripts encoding aggregation-prone proteins changes with immune load, and found a significant positive association with immune load for *APP, SQSTM1, TARDBP and MAPT* (**Fig. 4F**). These findings suggest that increased immune load is accompanied by enhanced expression of transcripts implicated in protein aggregation.

Next, we compared genes identified through linear regression with a high intercept (>4.0), representing transcripts with high predicted expression levels in the absence of immune burden, with genes identified by differential expression analysis between samples without detectable immune cells and those with low immune load. This approach identified genes whose expression patterns precede or accompany early immune infiltration and may represent pre-existing tissue states, susceptibility factors, or early molecular responses associated with immune infiltration. Using this approach, we identified 25 overlapping genes, including *ACTA2, ACTC1, ANKRD1, BIN1, DES, EEF1A1, EEF1G, FLII, HSPA8, HSPB1, HSPB8, MAP4, MAP7D1, MUSTN1, MYO18B, NFE2L1, PABPC4, PLEC, RYR1, SLN, STBD1, SVIL, SYNM, SYNPO2, and ZNF106* (**Fig. 4G**). These genes are enriched for functions related to muscle structure, contractile machinery, cytoskeletal organization, and proteostasis, consistent with a muscle-intrinsic remodeling state associated with early immune infiltration. Increased expression of *NFE2L1*, a regulator of proteasome homeostasis, may reflect elevated protein quality-control demands and enhanced processing of proteins, potentially contributing to antigen generation and immune presentation. Notably, several genes, including *NFE2L1*, *SYNPO2*, and *MYO18B*, were previously identified in regenerative muscle populations, suggesting that early regenerative states may precede or accompany immune-associated remodeling.

### Spatial niche analysis identifies an interferon-driven inflammatory microenvironment

Next, we sought to identify recurrent spatial niches in an unbiased manner, without imposing prior assumptions regarding neighborhood cellular composition. Using CellCharter [9], we defined a three-hop neighborhood expression embedding surrounding each myofiber and generated neighborhood embeddings capturing the local transcript signature of the myofiber microenvironment (**Fig. 5A**). Unsupervised Leiden clustering of these embeddings identified six recurrent myofiber-associated spatial niches (**Fig.5B**).

**Fig. 5:**
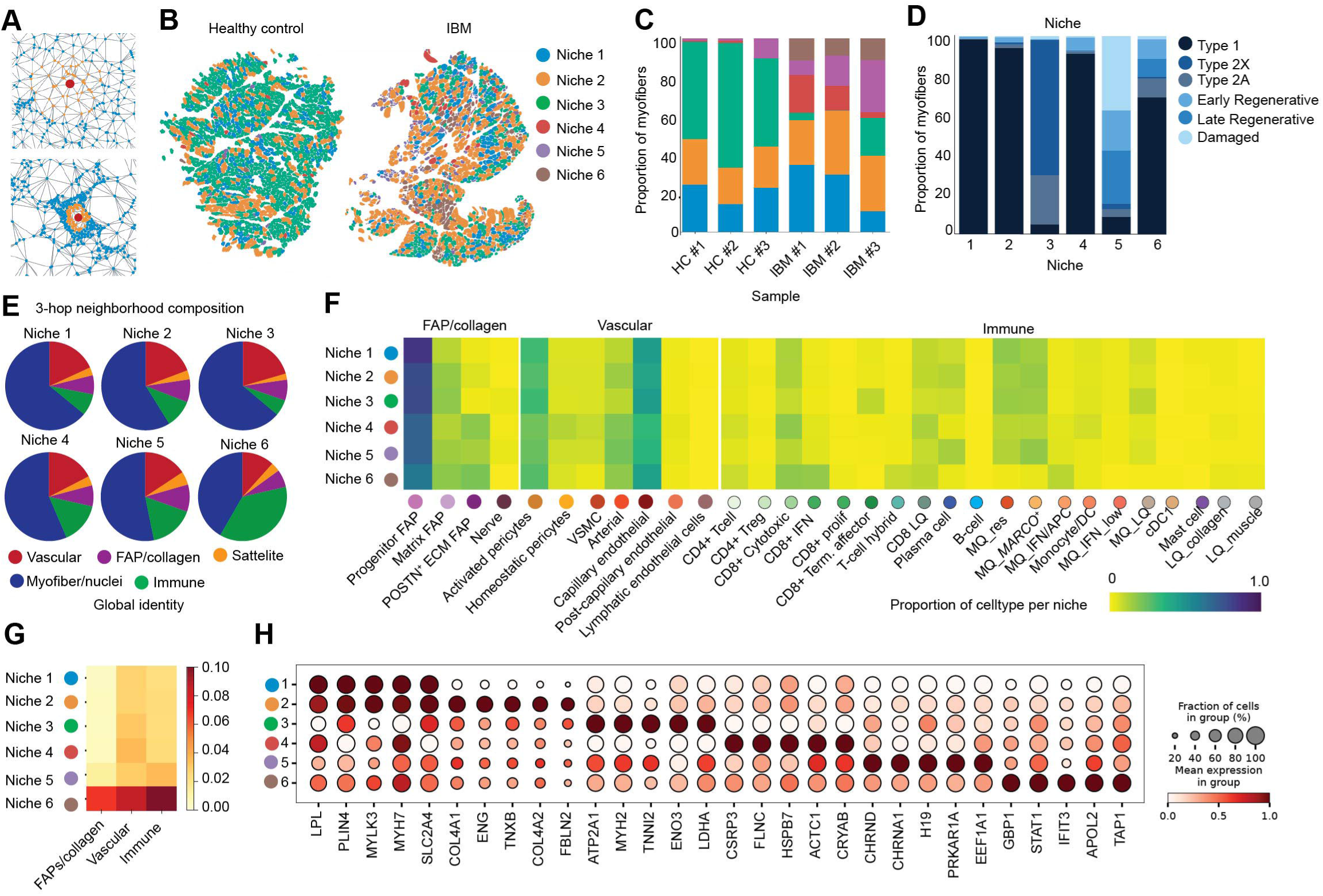
Spatial niche analysis identifies interferon-driven inflammatory microenvironments in IBM. **A)** Schematic overview of the CellCharter workflow. A three-hop neighborhood (orange) was defined around each myofiber (red dot), and neighborhood embeddings were generated to capture the cellular composition and transcriptomic context of the local microenvironment. **B)** Spatial maps of representative healthy control (HC) and IBM biopsies showing CellCharter-derived myofiber-associated spatial niches. Each myofiber is colored according to its assigned niche. **C)** Stacked bar plot showing the relative abundance of spatial niches across HC and IBM biopsies. **D)** Stacked bar plot showing the myofiber subtype composition of each spatial niche. **E)** Pie charts showing the broad cellular composition of the three-hop neighborhood surrounding myofibers assigned to each spatial niche, illustrating differences in the relative abundance of vascular, FAP, immune, and satellite cell compartments. **F)** Heatmap displaying the relative abundance of individual mononuclear cell populations within each spatial niche. **G)** Heatmap showing interferon (IFN) activation scores across spatial niches. **H)** Dot plot showing differential gene expression across for myofibers spatial niches.

Niches 1–3 were conserved across healthy control and IBM samples, whereas Niches 4–6 were preferentially expanded in IBM tissue, indicating the emergence of disease-associated spatial states (**Fig. 5C**). Niches 1 and 2 were defined by subtle differences in myofiber subtype distribution (**Fig. 5D**) and mononuclear cell composition, including differences in both the relative abundance of major cell type groups (**Fig.5E**), and the distribution of individual subtypes within each cell type group across niches (**Fig. 5F**). Niche 3 exhibited a more heterogeneous myofiber composition, predominantly comprising type 2 fibers and showing enrichment of terminal effector CD8+ T cells.

Niches 4–6 were composed predominantly of IBM-derived myofibers and differed in both myofiber subtype composition and the cellular composition of their three-hop neighborhoods. Niche 4 was enriched for type 1 myofibers, whereas Niche 5 was characterized by damaged, early and late regenerating myofibers, together with enrichment of plasma cells. Niche 6 represented a distinct IBM-associated spatial state containing type 1, damaged, and early and late regenerating myofibers, but was distinguished by a relative absence of homeostatic macrophage populations observed in Niches 1–5 together with the enrichment of IFN-responsive CD8+ T cells and IFN-stimulated antigen-presenting macrophages.

To determine whether these spatial niches reflected differences in cellular state in addition to differences in cellular composition, we compared the transcriptional profiles of each mononuclear cell type across niches. Differential expression analysis within individual mononuclear cell types revealed only modest transcriptional differences across Niches 1–5. In contrast, cells residing in Niche 6 consistently upregulated interferon genes, indicating a shared IFN-activated state across multiple mononuclear cell populations (**Supplement Fig. 8**). To further define the nature of this interferon response, we calculated pan-interferon, type I interferon, and type II interferon activation scores (**Supplement Fig. 8**). Across mononuclear immune populations, the interferon-associated state was predominantly driven by type II interferon signaling. Consistent with this, cells residing in Niche 6 displayed increased pan-interferon and type II interferon stimulated scores, indicating a coordinated type II interferon-associated inflammatory state across multiple mononucleated populations (**Fig. 5G**).

We next examined whether myofibers occupying the IBM-enriched niches (Niches 4–6) displayed distinct transcriptional states. Niche 4 was characterized by predominantly type 1 myofibers within a moderately inflammatory environment, marked by enrichment of cytotoxic CD8+ T cells and an associated moderate IFN-environment state. This suggests a spatial environment in which relatively preserved myofibers are exposed to moderate T-cell-associated signaling.

In contrast, Niche 5 was distinguished by the highest abundance of damaged and regenerating myofiber subtypes, together with enrichment of plasma cells and resident macrophage populations, suggesting a tissue injury and repair-associated environment. Niche 5 myofibers exhibited enrichment of neuromuscular junction-associated genes, including acetylcholine receptor components, together with cellular stress-associated programs (**Fig5.H**). Notably, these transcriptional changes were observed across fiber types and were not solely explained by the increased abundance of damaged or regenerating fibers within this niche (**Supplement Fig. 9**). Despite increased immune representation, this niche exhibited less pronounced IFN activation compared with Niche 4 and 6, coinciding with a distinct immune composition characterized by plasma cells and resident macrophages rather than (IFN-associated) cytotoxic T-cell populations. Consistent with the presence of plasma cells in Niche 5, as well as in Niche 2 and Niche 3 where plasma cells were also detected, myofibers in these niches showed increased immunoglobulin transcript abundance, including *IGHG1* and *IGHG4*, indicating a localized antibody-associated immune environment (**Supplementary Fig. 10**).

Niche 6 represented a distinct immune-dominant inflammatory state. Although containing a mixture of type 1, damaged, and regenerating myofibers, this niche was characterized by the highest immune cell abundance and strongest IFN activation across immune populations. In particular, Niche 6 was enriched for IFN-responsive CD8+ T cells and IFN-stimulated antigen-presenting macrophages, while showing reduced representation of resident macrophage populations. Transcripts upregulated in myofiber Niche 6 included IFN-responsive genes such as *GBP1, STAT1, IFIT3, APOL2*, and *TAP1*, corroborating the linear regression analysis results that identified an association between increased immune burden and IFN activation. Together, these features identify Niche 6 as a highly inflamed microenvironment characterized by coordinated immune activation and IFN-associated signaling. Consistent with this immune-enriched environment, Niche 6 myofibers displayed the highest immune cell abundance within a 200-pixel radius, suggesting that the spatial proximity of immune cells may contribute to the transcriptional changes associated with immune pressure (**Supplementary Fig. 11**). Together, these features identify Niche 6 as a highly inflamed microenvironment characterized by coordinated immune activation and IFN-associated signaling.

## Discussion

In this study, we used high-resolution spatial transcriptomics to define the cellular and spatial architecture underlying disease pathology of IBM muscle. By integrating complementary Xenium gene panels, we simultaneously resolved myofiber subtypes, immune cell composition, and the spatial organization of immune populations within intact muscle tissue. We identified IFN-responsive CD8⁺ T cells as the predominant expanded T-cell population in IBM, together with IFN-stimulated, antigen-presentation-competent macrophages, defining a spatially organized inflammatory microenvironment. Additionally, our spatial analysis reveals a complex immune repertoire that extends beyond the classical T-cell-centric view of IBM pathology. The immune landscape within IBM muscle tissue included multiple immune populations, including dendritic cells, B cells, and plasma cells. Importantly, immune infiltration into myofibers was not restricted to cytotoxic T cells but included multiple immune populations, indicating that myofiber invasion represents a broader multicellular process than previously recognized. Plasma cells were found throughout the biopsy but showed localized spatial enrichment, forming distinct plasma cell neighborhoods associated with increased immunoglobulin transcript abundance, including *IGHG1* and *IGHG4*, consistent with localized antibody-producing activity. Whether these plasma cells contribute directly to antigen recognition, immune amplification, or instead represent a response to chronic inflammation remains to be studied.

A central unresolved question in IBM is how the two defining pathological features of the disease— accumulation of protein aggregates and chronic inflammation—are mechanistically connected. Protein aggregates containing proteins such as APP, SQSTM1 (p62), TARDBP, and MAPT are characteristic hallmarks of IBM muscle, yet it remains unclear whether they actively initiate immune recruitment, represent a consequence of inflammatory stress, or reflect a parallel disease process. In our spatial analysis, aggregation-associated transcriptional programs were detected predominantly in early and late regenerating, as well as damaged myofibers and showed a significant positive association with increased immune load. This study provides the first spatial evidence linking aggregation-associated transcriptional programs in IBM myofibers with immune cell accumulation, indicating that aggregation-associated transcriptional programs and increased immune load could represent coupled features of disease pathology.

In addition to aggregation-associated transcripts, immune-enriched myofibers displayed activation of interferon type I-and type II and antigen presentation-related programs. Among the transcripts positively associated with increasing immune burden was *WARS* (tryptophanyl-tRNA synthetase). Aminoacyl-tRNA synthetases have been implicated in autoimmune myopathies, most notably through Jo-1 autoantibody responses against specific synthetases such as histidyl-tRNA synthetase (HARS) in antisynthetase syndrome [11–13]. Although *WARS* belongs to this protein family, a role for *WARS* in IBM has not been extensively described. Its association with increasing immune burden in our dataset raises the possibility that interferon-associated regulation of aminoacyl-tRNA synthetases may represent a shared molecular feature linking IBM with other autoimmune myopathies. However, whether *WARS* contributes functionally to immune activation in IBM, or instead reflects a broader interferon-induced myofiber state, remains to be determined.

The presence of baseline myofiber expression programs that overlap with those activated during early immune infiltration suggests that distinct myofiber states may represent early events that precede or accompany immune recruitment in IBM. These genes were enriched for muscle structural organization, contractile function, cytoskeletal remodeling, and proteostasis, indicating that immune-associated changes occur within a context of altered intrinsic myofiber biology. Among these genes, *NFE2L1* is of particular interest due to its central role in maintaining proteostasis through regulation of proteasome homeostasis and its ability to modulate protein quality-control pathways, including *SQSTM1* (p62)-mediated aggregate clearance [14–16]. Increased *NFE2L1* expression in regenerative myofibers and myofibers that start to be surrounded by immune cells may therefore reflect increased proteostatic demands associated with altered protein turnover and potential aggregate handling. Such changes could influence the availability of intracellular proteins for processing and peptide presentation, providing a potential link between myofiber-intrinsic proteostasis alterations and immune presentation.

Interestingly, *NFE2L1* has also been implicated in cancer, where expression has been associated with immune evasion mechanisms, including modulation of PD-L1 (*CD274*) expression [17, 18]. PD-L1 expression can suppress CD8⁺ T-cell activity and contribute to dysfunctional or exhausted T-cell states. Notably, *PD-L1* expression itself was not enriched in immune-associated myofibers in our dataset. However, the enrichment of *NFE2L1*, a regulator that has been linked to PD-L1-mediated immune evasion in cancer contexts, is of interest given the expansion of IFN-associated and partially exhausted CD8+ T-cell populations observed in IBM. Whether *NFE2L1* contributes to the CD8 T-cell activation/exhaustion in IBM through regulation of checkpoint-related pathways, co-stimulatory or inhibitory immune signals remains to be determined.

Our spatial niche analysis revealed a distinct IFN-dominant inflammatory microenvironment characterized by cell-type-specific interferon responses. While myofibers within this environment exhibited activation of both type I and type II interferon programs, together with increased expression of antigen presentation machinery, surrounding mononuclear immune cells showed a predominantly type II interferon signature, highlighting a coordinated but distinct interferon response between immune cells and myofibers.

Although circulating autoantibodies, most notably anti-cN1A antibodies, have been identified in a subset of IBM patients [19, 20], the specific muscle-derived antigens that drive autoreactive T-cell responses remain unknown. In our spatial analysis, immune-enriched myofibers displayed increased expression of antigen presentation-associated transcripts, including components of both the MHC class I and MHC class II antigen presentation machinery. In contrast, the surrounding mononucleated immune populations, particularly IFN-stimulated CD8⁺ T cells and antigen-presentation-competent macrophages, were predominantly characterized by type II interferon-associated signatures. Together, these findings support a model in which myofiber-intrinsic alterations in protein homeostasis may represent an early component of immune niche formation in IBM. Increased proteostatic demand, reflected by enrichment of genes such as *NFE2L1*, may influence proteasome activity and the processing of intracellular proteins, potentially affecting MHC class I antigen presentation. In the context of IBM, where CD8⁺ T cells represent the dominant expanded lymphocyte population, enhanced presentation of muscle-derived peptides by myofibers may contribute to recruitment and/or retention of CD8⁺ T cells surrounding affected myofibers. The sustained presence and activation of these cells may promote local IFN-type II production, further enhancing inflammatory signaling within the tissue. This may establish a feed-forward inflammatory environment in which type I interferon-associated responses within myofibers coexist with type II interferon-driven activation from surrounding immune cells.

Our findings suggest that IBM pathology may be sustained by a reciprocal interaction between immune activation and myofiber-intrinsic alterations. Although selective targeting of *KLRG-1*⁺ IFN-responsive CD8⁺ T cells represents a potential strategy to modulate a major cytotoxic immune component in IBM, our spatial analysis indicates that immune repertoire are characterized by broader interactions involving macrophages, dendritic cells, B cells, plasma cells, and myofibers with an increased capacity for antigen presentation. Therefore, therapeutic approaches directed solely at a single immune effector population may not fully disrupt disease-associated niches if antigen presentation and inflammatory signaling from the tissue environment persist. Targeting interferon-driven inflammation, including modulation of type I interferon-associated responses in myofibers or type II interferon-associated activation in immune cells, represents a potential strategy to interrupt these inflammatory circuits. Furthermore, the enrichment of proteostasis regulators such as *NFE2L1* highlights the possibility that modulating protein quality-control pathways may influence both aggregate-associated pathology and immune recognition. Together, these findings provide a rationale for exploring therapeutic strategies that target both immune activation and muscle-intrinsic disease mechanisms in IBM.

## Materials and Methods

### Human muscle biopsies

Fresh frozen muscle biopsies from three IBM patients, fulfilling the ENMC 2024 diagnostic criteria [21], and three healthy controls were taken and processed for spatial transcriptomic profiling. An overview of muscle type used is listed in Table 1. All human material used was derived from biobanks for which licenses and informed consent had been previously obtained.

**Table 1:**

| Sample | Condition | Muscle type |
| --- | --- | --- |
| 1 | IBM | Tibialis anterior |
| 2 | IBM | Tibialis anterior |
| 3 | HC | Gastrocnemius lateralis |
| 4 | HC | Vastus lateralis |
| 5 | HC | Gastrocnemius lateralis |
| 6 | IBM | Vastus lateralis |

### Spatial Transcriptomics

Spatial transcriptomic profiling was performed using the 10x Genomics Xenium platform. 10 µm serial sections were placed on Xenium slides and analyzed using two complementary probe panels: 1) Xenium Human Pan-Tissue & Pathway Panel (5K panel); 2) Custom-designed 480-gene panel enriched for skeletal muscle biology, inflammatory pathways, antigen presentation, and IBM-associated transcripts (480 panel). Xenium data was previously generated according to manufacturer’s protocols.

### Data preprocessing

Raw Xenium output was converted into a SpatialData object and stored in the Zarr format using the SpatialData framework. Myofibers were segmented from the morphology images using the Cellpose Cyto3 deep learning model as implemented in the Sparrow framework (v.0.0.1). Mononuclear cell nuclei were segmented using the default DAPI-based nuclei segmentation provided by Xenium Ranger, with the cell expansion buffer set to 0 µm. Transcript were allocated to segmentation mask and normalized to cell size using Sparrow’s tb.allocate() function.

### Fiber integration

Serial sections from the 5K and 480-gene panels were aligned separately for each sample and visually distinct muscle lobe using manually defined landmarks in napari-spatialdata (v0.6.0). At least three corresponding landmarks per lobe were used to calculate rigid transformations with SpatialData (v0.5.0), aligning the 5K fiber segmentations to the 480-gene sections. Fibers were matched based on overlap of the transformed segmentation masks and Pearson correlation of expression across 198 overlapping genes between panels. MYH1, MYH2, and LDB3 were excluded from the correlation analysis because of fiber-type-specific expression or differences in probe sensitivity between panels. Fiber matches were retained when Pearson’s *r* > 0.6, with the threshold determined from the distribution of correlations among high-confidence shape-based matches (>60% overlap). All matches were subsequently verified by visual inspection.

### Data Quality Control, clustering and annotation

Transcriptomic profiles were analyzed using a graph-based clustering workflow implemented in Scanpy. A k-nearest neighbor graph was constructed based on the 50 nearest neighbors using the first 10 principal components to define transcriptional relationships among observations. Unsupervised clustering was performed using the Leiden community detection algorithm with a resolution parameter of 1.0 to identify transcriptionally distinct clusters. UMAP dimensionality reduction was subsequently applied to visualize the transcriptional structure and distribution of identified clusters in a low-dimensional embedding space. Clusters with low transcript counts (<200) and lack of marker gene expression where removed for the dataset or labelled as ‘low quality’ in downstream analysis.

### Infiltrated immune cells into myofiber

A custom Python script was developed to assign nuclei to predefined spatial regions based on segmentation-derived polygon masks. Segmentation boundary layers were transformed into a common global coordinate system using the SpatialData framework, and nuclei boundary geometries were similarly standardized prior to analysis. polygon masks were computationally eroded before evaluating spatial relationships. Each nucleus was tested for containment within individual spatial masks, and nuclei were assigned to a region if they were located within at one of the evaluated segmentation boundaries. A combined binary annotation was generated to identify nuclei associated with the defined spatial regions and was used for downstream spatial analyses

### Neighborhood analysis

Squidpy’s (v.1.6.5’s) spatial neighbors() function was used to generate a spatial neighborhood graph from the spatial coordinates of the centroids of all immune related segmented shapes in the Xenium datasets. Parameters used included coord type=’generic’ and n_neighs=50.

### Immune pixel radius

To quantify the number of immune cells in the neighborhood of myofibers, GeoPandas v1.1.2’s buffer() function was used to expand myofiber segmentation shapes by 200 pixels.

### Linear regression

For each gene, a linear regression model was fitted using SciPy v1.15.3s linregress() function to assess the association between gene expression (fiber size normalized and log-transformed counts) and the number of immune cells (log-transformed) in the fiber neighborhoods.

### Niche analysis

CellCharter v0.3.5 (Varrone et al., 2024) was used to define spatially aware clusters, or spatial niches, in the Xenium spatial transcriptomics data. CellCharter’s aggregate neighbors() function was used to compute a neighborhood embedding for each cell. The CellCharter implementation of GMMwas run on aGPUusing PyTorch v.2.4.0 following the CellCharter documentation. Finally, the spatial niches that were generated with CellCharter using trVAE and Leiden clustering were used in further analyses.

### IFN score

IFN signalling scores were calculated using the score_genes () function implemented in Scanpy, which calculates the average expression of signature genes normalized against a background gene set. Only genes detected within each dataset were included for score calculation. Three interferon-associated signatures were evaluated, including a pan-interferon response signature, a type I interferon (IFN-α/β)-associated antiviral response signature, and a type II interferon (IFN-γ)-associated inflammatory and antigen presentation signature. The pan-interferon signature consisted of *STAT1*, *IRF1*, *IFI35*, *WARS*, *TRAFD1*, *TAP1*, *TAP2*, *B2M*, and *IFITM1*. The type I interferon signature included *ISG15*, *IFIT1*, *IFIT2*, *IFIT3*, *MX1*, *MX2*, *OAS1*, *OAS2*, *OAS3*, *OASL*, *IFI6*, *IFI27*, *IFI44*, *IFI44L*, *RSAD2*, *BST2*, *IRF7*, *APOL1*, *APOL2*, *APOL3*, and *APOBEC3C*. The type II interferon signature consisted of *CXCL9*, *CXCL10*, *GBP1*, *GBP2*, *GBP5*, *IFI30*, *CIITA*, *HLA-DRA*, *HLA-DRB1*, *HLA-DPA1*, *HLA-DPB1*, *HLA-DQA1*, *CD74*, and *IDO1*.

## Author contribution

Conceptualization: SH, CA, LH, RA, HK, UB, AM, PS

Methodology: SH, CA, LH, UB, AM, PS

Investigation: SH, CA, UB, AM, PS

Visualization: SH, CA

Funding acquisition: HK (ERN EURO NMD), PS (ERN EURO NMD), SH (EMBO_ALT92_2025)

Project administration: PS

Supervision: UB, AM, PS

Writing – original draft: SH, CA, UB, AM, PS

Writing – review & editing: SH, CA, RA, HK, UB, AM, PS

## Data and code availability

Analysis code generated during this study will be made publicly available upon publication of this manuscript in an appropriate public repository. Accession numbers and repository links will be provided at the time of publication.

## Acknowledgements

We like to thank the Leiden Genome Technology Center (LGTC, Leiden University Medical Center) for spatial transcriptomics technical support. We also thank all members of the NMD Biomarker group and Mahfouz groups for scientific discussions and/or technical support. We also thank Dr. Maaike van Putten (Leiden University Medical Center) for scientific discussions. This work was carried out within the framework of the European Reference Network for Neuromuscular Diseases (ERN EURO NMD).

## Declaration of interests

all authors declare no competing interest.

## Inventory of Supplement Information

### Supplementary Figures

**Sup. Fig. 1:** Expression patterns of immunoglobin genes surrounding plasma cells

**Sup. Fig. 2:** KLRG-1 positivity across T-cell populations

**Sup. Fig. 3:** Expression patterns of T-cell and macrophage markers in cells annotated as hybrid T-cells

**Sup. Fig. 4:** Number of immune cells normalized to 100 myofibers per dataset

**Sup. Fig. 5:** KLRG-1 ligand expression on cDC1 cells

**Sup. Fig. 6:** Illustration of 200 pixel radius surrounding myofibers

**Sup. Fig. 7:** Identified cell surface genes expressed in IFN-responsive CD8+ T-cells

**Sup. Fig. 8:** Differentially expressed genes in mononucleated cell populations across niches and IFN-scores across niches

**Sup. Fig. 9:** Differentially expressed genes in myofiber populations across niches

**Sup. Fig. 10:** Plasma cell related transcript expression in myofibers across niches

**Sup. Fig. 11:** Total immune cells per niche

### Supplementary Tables

**Sup. Table.1:** Xenium gene panels

**Sup. Table.2:** Differentially expressed genes of myofiber populations

**Sup. Table.3:** Differentially expressed genes of immune populations

**Sup. Table.4:** Candidate genes identified in IFN-responsive CD8+ T-cells

**Sup. Table.5:** Linear regression results all myofibers

**Sup. Table.6:** Linear regression results per myofiber subtype

## Notes

### Competing Interest Statement

The authors have declared no competing interest.

### Author Declarations

Ethics committee/IRB of the Leiden University Medical Center gave ethical approval for this work

### Summary of Updates

The first submitted version did not include main figures of the manuscript

## References

1. Dimachkie, M.M. and R.J. Barohn, Inclusion body myositis. Curr Neurol Neurosci Rep, 2013. 13(1): p. 321.

2. Salajegheh, M.K. and A.A. Amato, Idiopathic Inflammatory Myopathies. Continuum (Minneap Minn), 2025. 31(5): p. 1385–1408.

3. Goyal, N.A., et al., Immunophenotyping of Inclusion Body Myositis Blood T and NK Cells. Neurology, 2022. 98(13): p. e1374–e1383.

4. Badrising, U.A., et al., Optimism in inclusion body myositis: a double-blind randomised controlled phase III trial investigating the effect of sirolimus on disease progression in patients with IBM as measured by the IBM Functional Rating Scale. Clin Exp Rheumatol, 2025. 43(2): p. 316–325.

5. Ruxolitinib Inclusion Body Myositis.

6. Wischnewski, S., et al., Cell type mapping of inflammatory muscle diseases highlights selective myofiber vulnerability in inclusion body myositis. Nat Aging, 2024. 4(7): p. 969–983.

7. Pinal-Fernandez, I., et al., Spatial transcriptomics reveals mechanism of autoimmunity driven by internalized autoantibodies. medRxiv, 2026.

8. Liang, R., et al., TIGIT promotes CD8(+)T cells exhaustion and predicts poor prognosis of colorectal cancer. Cancer Immunol Immunother, 2021. 70(10): p. 2781–2793.

9. Greenberg, S.A., et al., Highly differentiated cytotoxic T cells in inclusion body myositis. Brain, 2019. 142(9): p. 2590–2604.

10. Kirou, R.A., et al., Activated Dendritic Cell Subsets Characterize Muscle of Inclusion Body Myositis Patients and Correlate with KLRG1+ and TBX21+ CD8+ T cells. medRxiv, 2025.

11. Park, S.G., P. Schimmel, and S. Kim, Aminoacyl tRNA synthetases and their connections to disease. Proc Natl Acad Sci U S A, 2008. 105(32): p. 11043–9.

12. Muro, Y., et al., Two novel anti-aminoacyl tRNA synthetase antibodies: Autoantibodies against cysteinyl-tRNA synthetase and valyl-tRNA synthetase. Autoimmun Rev, 2022. 21(12): p. 103204.

13. Robinson, D. and B. Scholz, The antisynthetase syndrome. Proc (Bayl Univ Med Cent), 2020. 33(3): p. 401–403.

14. Chandran, A., H.J. Oliver, and J.C. Rochet, Role of NFE2L1 in the Regulation of Proteostasis: Implications for Aging and Neurodegenerative Diseases. Biology (Basel), 2023. 12(9).

15. Hatanaka, A., et al., The transcription factor NRF1 (NFE2L1) activates aggrephagy by inducing p62 and GABARAPL1 after proteasome inhibition to maintain proteostasis. Sci Rep, 2023. 13(1): p. 14405.

16. Smahelova, Z., et al., Investigating NFE2L1 activators for targeted protein aggregate clearance: a follow-up study. RSC Med Chem, 2025. 16(12): p. 6397–6411.

17. Shi, C., et al., Identifying a locus in super-enhancer and its resident NFE2L1/MAFG as transcriptional factors that drive PD-L1 expression and immune evasion. Oncogenesis, 2023. 12(1): p. 56.

18. Kankanamge, L.P., et al., Nrf1 coordinates proteasome activity and autophagy to maintain cardiac proteostasis. Commun Biol, 2026. 9(1).

19. Patrikiou, E., C. Liaskos, and D.P. Bogdanos, On the role of anti-cN1A antibodies in sporadic inclusion body myositis and beyond: a challenging task full of surprises. Reumatologia, 2023. 61(6): p. 411–413.

20. Herbert, M.K., et al., Disease specificity of autoantibodies to cytosolic 5’-nucleotidase 1A in sporadic inclusion body myositis versus known autoimmune diseases. Ann Rheum Dis, 2016. 75(4): p. 696–701.

21. Lilleker, J.B., et al., 272nd ENMC international workshop: 10 Years of progress - revision of the ENMC 2013 diagnostic criteria for inclusion body myositis and clinical trial readiness. 16-18 June 2023, Hoofddorp, The Netherlands. Neuromuscul Disord, 2024. 37: p. 36–51.

