## Supplement Figures for "High-Resolution Spatial Transcriptomics Reveals Interferon-Associated Immune Niches and Antigen Presentation Programs in Inclusion Body Myositis"

**Sup. Fig. 1**

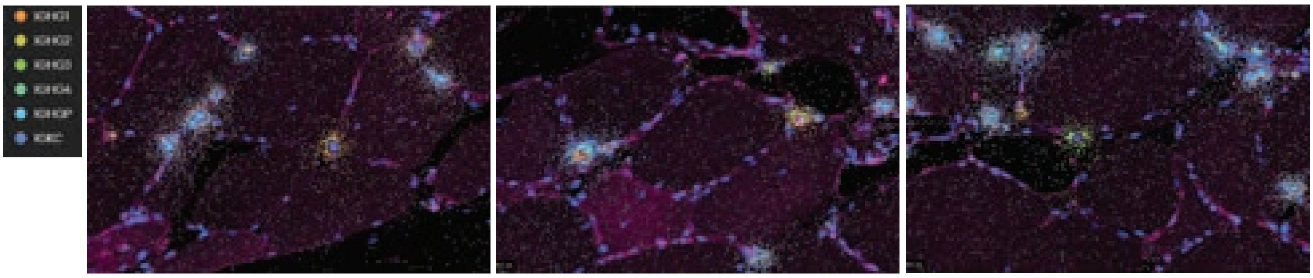

**Sup. Fig. 2**

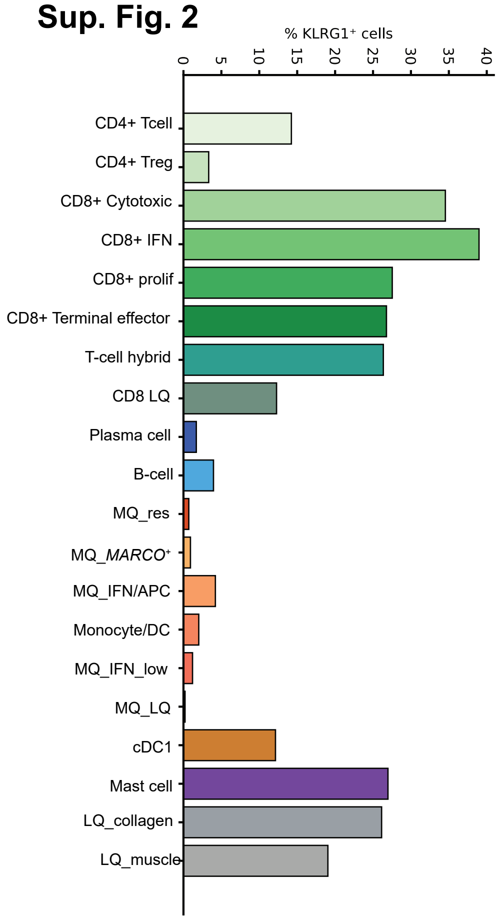
% KLRG1⁺ cells
0
5
10
15
20
25
30
35
40
CD4+ Tcell
CD4+ Treg
CD8+ Cytotoxic
CD8+ IFN
CD8+ prolif
CD8+ Terminal effector
T-cell hybrid
CD8 LQ
Plasma cell
B-cell
MQ_res
MQ_MARCO⁺
MQ_IFN/APC
Monocyte/DC
MQ_IFN_low
MQ_LQ
cDC1
Mast cell
LQ_collagen
LQ_muscle

### Sup. Fig. 3

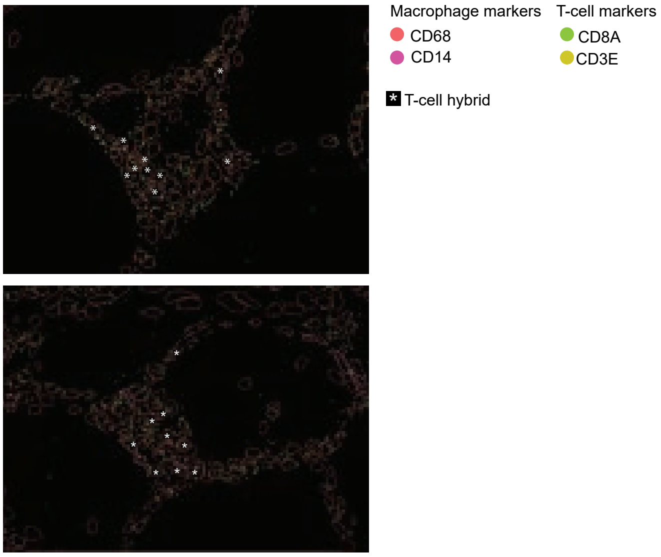

Macrophage markers
CD68
CD14
T-cell markers
CD8A
CD3E
T-cell hybrid

#### Sup. Fig. 4

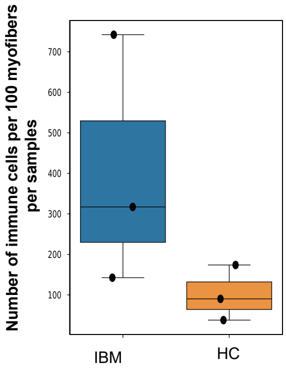
Number of immune cells per 100 myofibers per samples
700
600
500
400
300
200
100
IBM
HC

**Sup. Fig. 5**

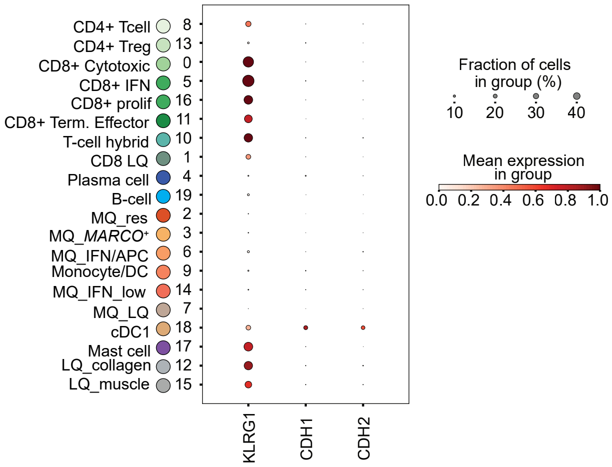

CD4+ Tcell 8
CD4+ Treg 13
CD8+ Cytotoxic 0
CD8+ IFN 5
CD8+ prolif 16
CD8+ Term. Effector 11
T-cell hybrid 10
CD8 LQ 1
Plasma cell 4
B-cell 19
MQ_res 2
MQ_MARCO+ 3
MQ_IFN/APC 6
Monocyte/DC 9
MQ_IFN_low 14
MQ_LQ 7
cDC1 18
Mast cell 17
LQ_collagen 12
LQ_muscle 15
KLRG1
CDH1
CDH2
Fraction of cells in group (%)
10 20 30 40
Mean expression in group
0.0 0.2 0.4 0.6 0.8 1.0

**Sup. Fig. 6**

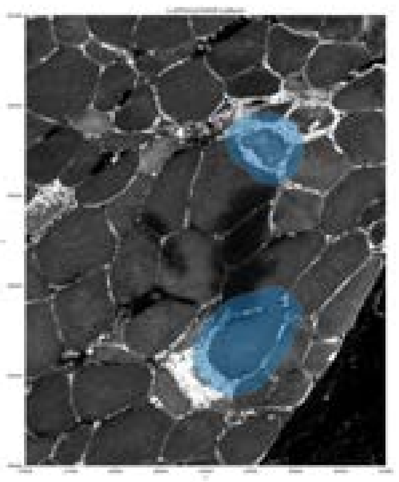

**Sup. Fig. 7**

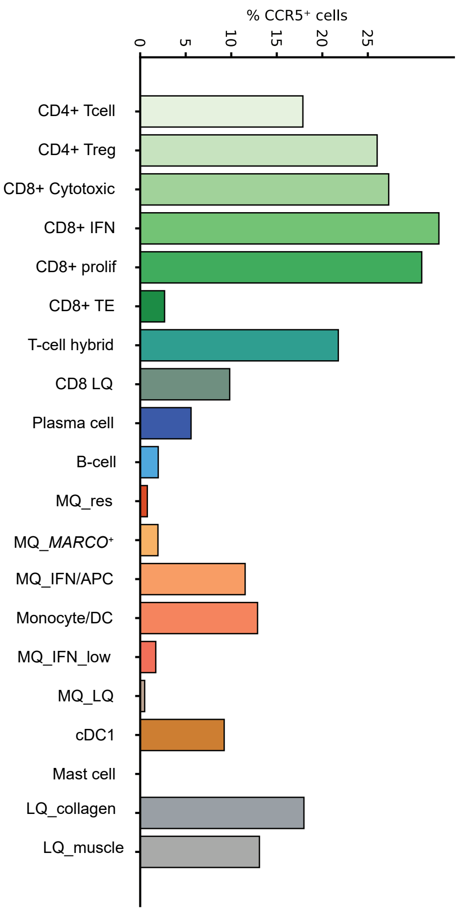
% CCR5⁺ cells
0
5
10
15
20
25
CD4+ Tcell
CD4+ Treg
CD8+ Cytotoxic
CD8+ IFN
CD8+ prolif
CD8+ TE
T-cell hybrid
CD8 LQ
Plasma cell
B-cell
MQ_res
MQ_MARCO⁺
MQ_IFN/APC
Monocyte/DC
MQ_IFN_low
MQ_LQ
cDC1
Mast cell
LQ_collagen
LQ_muscle

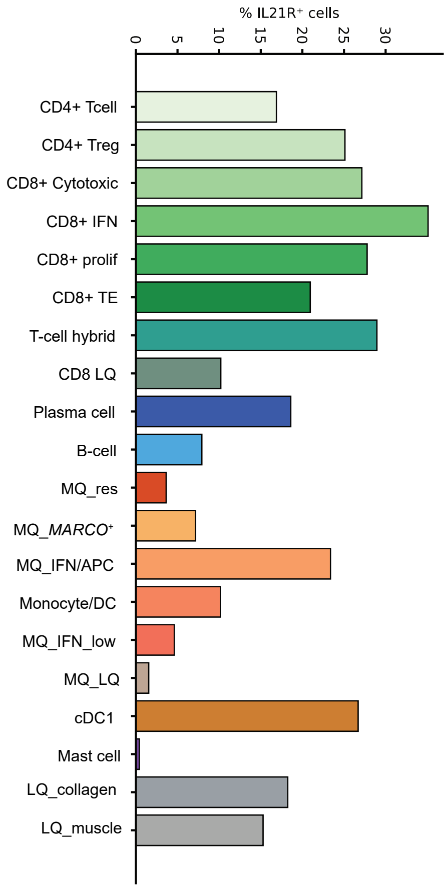
% IL21R⁺ cells
0
5
10
15
20
25
30
CD4+ Tcell
CD4+ Treg
CD8+ Cytotoxic
CD8+ IFN
CD8+ prolif
CD8+ TE
T-cell hybrid
CD8 LQ
Plasma cell
B-cell
MQ_res
MQ_MARCO⁺
MQ_IFN/APC
Monocyte/DC
MQ_IFN_low
MQ_LQ
cDC1
Mast cell
LQ_collagen
LQ_muscle

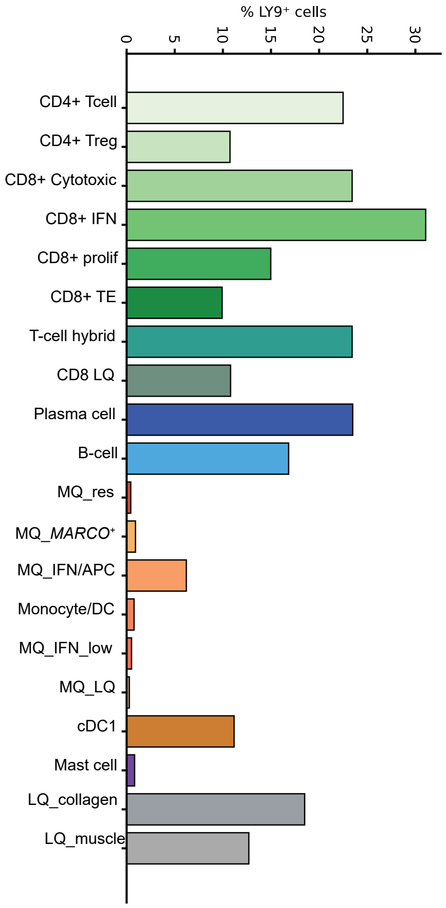
% LY9⁺ cells
0
5
10
15
20
25
30
CD4+ Tcell
CD4+ Treg
CD8+ Cytotoxic
CD8+ IFN
CD8+ prolif
CD8+ TE
T-cell hybrid
CD8 LQ
Plasma cell
B-cell
MQ_res
MQ_MARCO⁺
MQ_IFN/APC
Monocyte/DC
MQ_IFN_low
MQ_LQ
cDC1
Mast cell
LQ_collagen
LQ_muscle

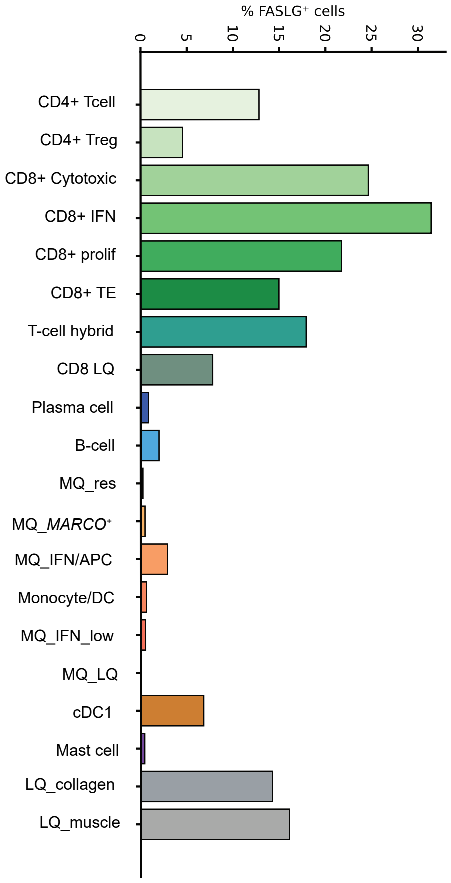
% FASLG⁺ cells
0
5
10
15
20
25
30
CD4+ Tcell
CD4+ Treg
CD8+ Cytotoxic
CD8+ IFN
CD8+ prolif
CD8+ TE
T-cell hybrid
CD8 LQ
Plasma cell
B-cell
MQ_res
MQ_MARCO⁺
MQ_IFN/APC
Monocyte/DC
MQ_IFN_low
MQ_LQ
cDC1
Mast cell
LQ_collagen
LQ_muscle

### Sup. Fig. 8

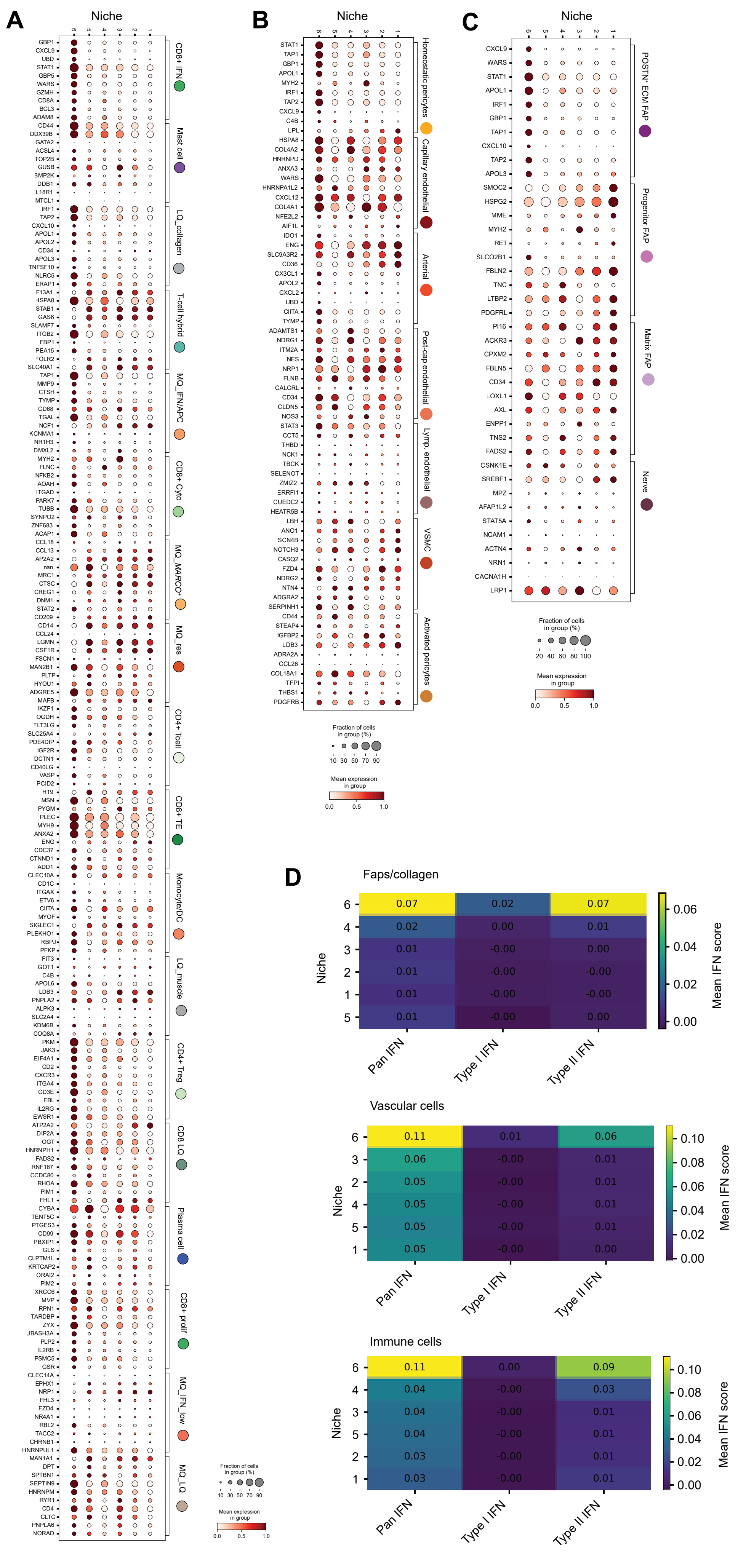

A
Niche
CD8+ IFN
Mast cell
LQ_collagen
T-cell hybrid
MQ_IFN/APC
CD8+ Cyto
MQ_MARCO+
MQ_res
CD4+ Tcell
CD8+ TE
Monocyte/DC
LQ_muscle
CD4+ Treg
CD8 LQ
Plasma cell
CD8+ prolif
MQ_IFN_low
MQ_LQ
Fraction of cells in group (%)
10 30 50 70 90
Mean expression in group
0.0 0.5 1.0
B
Niche
Homeostatic pericytes
Capillary endothelial
Arterial
Post-cap endothelial
Lymp. endothelial
VSMC
Activated pericytes
Fraction of cells in group (%)
10 30 50 70 90
Mean expression in group
0.0 0.5 1.0
C
Niche
POSTN+ ECM FAP
Progenitor FAP
Matrix FAP
Nerve
Fraction of cells in group (%)
20 40 60 80 100
Mean expression in group
0.0 0.5 1.0
D
Faps/collagen
Niche
6 0.07 0.02 0.07
4 0.02 0.00 0.01
3 0.01 -0.00 0.00
2 0.01 -0.00 -0.00
1 0.01 -0.00 -0.00
5 0.01 -0.00 0.00
Pan IFN
Type I IFN
Type II IFN
Mean IFN score
0.06 0.04 0.02 0.00
Vascular cells
6 0.11 0.01 0.06
3 0.06 -0.00 0.01
2 0.05 -0.00 0.01
4 0.05 -0.00 0.01
5 0.05 -0.00 0.01
1 0.05 -0.00 0.00
Pan IFN
Type I IFN
Type II IFN
Mean IFN score
0.10 0.08 0.06 0.04 0.02 0.00
Immune cells
6 0.11 0.00 0.09
4 0.04 -0.00 0.03
3 0.04 -0.00 0.01
5 0.04 -0.00 0.01
2 0.03 -0.00 0.01
1 0.03 -0.00 0.01
Pan IFN
Type I IFN
Type II IFN
Mean IFN score
0.10 0.08 0.06 0.04 0.02 0.00

### Sup. Fig. 9

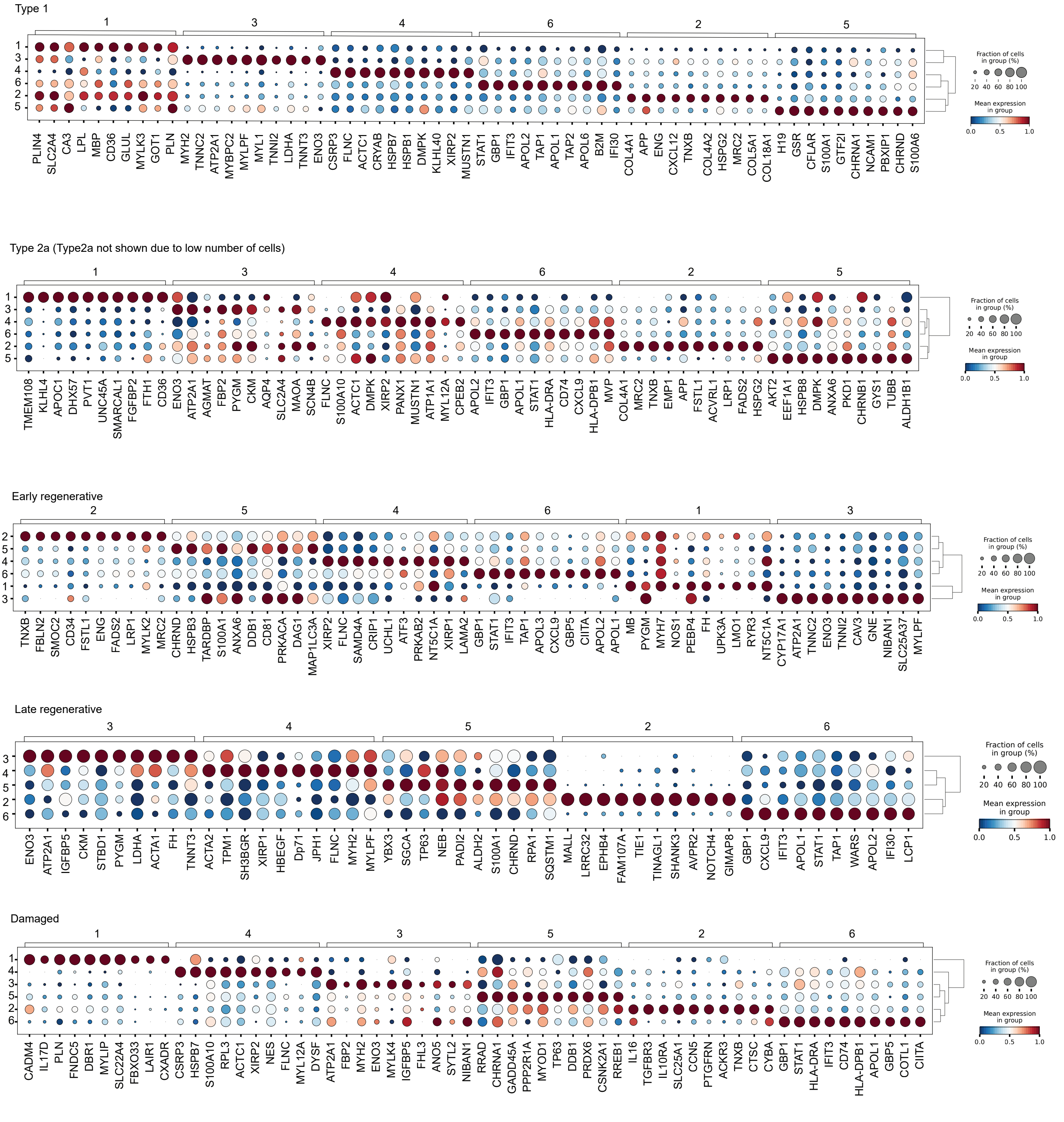
Type 1
1 3 4 6 2 5
PLIN4 SLC2A4 CA3 LPL MBP CD36 GLUL MYLK3 GOT1 PLN
MYH2 TNNC2 ATP2A1 MYBPC2 MYLPF MYL1 TNNI2 LDHA TNNT3 ENO3
CSRP3 FLNC ACTC1 CRYAB HSPB7 HSPB1 DMPK KLHL40 XIRP2 MUSTN1
STAT1 GBP1 IFIT3 APOL2 TAP1 APOL1 TAP2 APOL6 B2M IFI30
COL4A1 APP ENG CXCL12 TNXB COL4A2 HSPG2 MRC2 COL5A1 COL18A1
H19 GSR CFLAR S100A1 GTF2I CHRNA1 NCAM1 PBXIP1 CHRND S100A6
Type 2a (Type2a not shown due to low number of cells)
1 3 4 6 2 5
TMEM108 KLHL4 APOC1 DHX57 PVT1 UNC45A SMARCAL1 FGFBP2 FTH1 CD36
ENO3 ATP2A1 AGMAT FBP2 PYGM CKM AQP4 SLC2A4 MAOA SCN4B
FLNC S100A10 ACTC1 DMPK XIRP2 PANX1 MUSTN1 ATP1A1 MYL12A CPEB2
APOL2 IFIT3 GBP1 APOL1 STAT1 HLA-DRA CD74 CXCL9 HLA-DPB1 MVP
COL4A1 MRC2 TNXB EMP1 APP FSTL1 ACVRL1 LRP1 FADS2 HSPG2
AKT2 EEF1A1 HSPB8 DMPK ANXA6 PKD1 CHRNB1 GYS1 TUBB ALDH1B1
Early regenerative
2 5 4 6 1 3
TNXB FBLN2 SMOC2 CD34 FSTL1 ENG FADS2 LRP1 MYLK2 MRC2
CHRND HSPB3 TARDBP S100A1 ANXA6 DDB1 CD81 PRKACA DAG1 MAP1LC3A
XIRP2 FLNC SAMD4A CRIP1 UCHL1 ATF3 PRKAB2 NT5C1A XIRP1 LAMA2
GBP1 STAT1 IFIT3 TAP1 APOL3 CXCL9 GBP5 CIITA APOL2 APOL1
MB PYGM MYH7 NOS1 PEBP4 FH UPK3A LMO1 RYR3 NT5C1A
CYP17A1 ATP2A1 TNNC2 ENO3 TNNI2 CAV3 GNE NIBAN1 SLC25A37 MYLPF
Late regenerative
3 4 5 2 6
ENO3 ATP2A1 IGFBP5 CKM STBD1 PYGM LDHA ACTA1 FH TNNT3
ACTA2 TPM1 SH3BGR XIRP1 HBEGF Dp71 JPH1 FLNC MYH2 MYLPF
YBX3 SGCA TP63 NEB PADI2 ALDH2 S100A1 CHRND RPA1 SQSTM1
MALL LRRC32 EPHB4 FAM107A TIE1 TINAGL1 SHANK3 AVPR2 NOTCH4 GIMAP8
GBP1 CXCL9 IFIT3 APOL1 STAT1 TAP1 WARS APOL2 IFI30 LCP1
Damaged
1 4 3 5 2 6
CADM4 IL17D PLN FNDC5 DBR1 MYLIP SLC22A4 FBXO33 LAIR1 CXADR
CSRP3 HSPB7 S100A10 RPL3 ACTC1 XIRP2 NES FLNC MYL12A DYSF
ATP2A1 FBP2 MYH2 ENO3 MYLK4 IGFBP5 FHL3 ANO5 SYTL2 NIBAN1
RRAD CHRNA1 GADD45A PPP2R1A MYOD1 TP63 DDB1 PRDX6 CSNK2A1 RREB1
IL16 TGFBR3 IL10RA SLC25A1 CCN5 PTGFRN ACKR3 TNXB CTSC CYBA
GBP1 STAT1 HLA-DRA IFIT3 CD74 HLA-DPB1 APOL1 GBP5 COTL1 CIITA
Fraction of cells in group (%)
20 40 60 80 100
Mean expression in group
0.0 0.5 1.0

**Sup. Fig. 10**

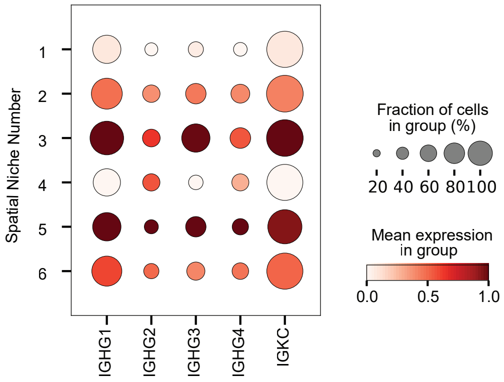

Spatial Niche Number
1
2
3
4
5
6
IGHG1
IGHG2
IGHG3
IGHG4
IGKC
Fraction of cells in group (%)
20 40 60 80 100
Mean expression in group
0.0 0.5 1.0

### Sup. Fig. 11

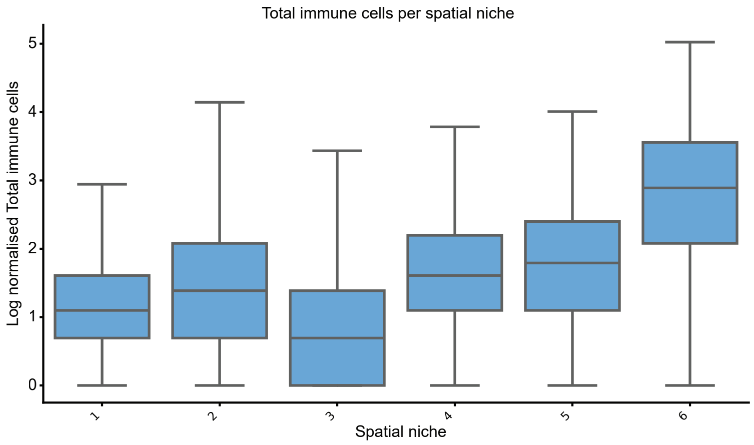
Total immune cells per spatial niche
Log normalised Total immune cells
0
1
2
3
4
5
1
2
3
4
5
6
Spatial niche
